# Patterns of use of mental health crisis-related services in the English NHS: a retrospective observational study

**DOI:** 10.64898/2026.01.08.25343254

**Authors:** Maria Ana Matias, Jessica L. Griffiths, Alan Simpson, Anam Bhutta, Andrew Grundy, Beverley Chipp, Jo Lomani, Jude Beng, Julian Edbrooke-Childs, Kylee Trevillion, Mark Holden, Martin Webber, Patrick Nyikavaranda, Rachel Rowan Olive, David P. Osborn, Rowena Jacobs, Sonia Johnson, Brynmor Lloyd-Evans

## Abstract

**Background:** Improving access to high-quality community-based crisis care and reducing inpatient admissions are key policy objectives in England, yet evidence on who uses crisis-related services and potential inequalities in access remains limited.

**Aims:** To examine socio-demographic patterns in the use of six crisis-related services in England: 1) Crisis phonelines; 2) Crisis cafés; 3) Crisis Resolution and Home Treatment teams (CRHTs); 4) Voluntary inpatient admissions; 5) Compulsory inpatient admissions under the Mental Health Act 1983 (MHA); and 6) conveyance to a Place of Safety (PoS) by police under section 136 of the MHA.

**Method:** We conducted a retrospective observational study using the Mental Health Services Data Set between 2021/22 and 2023/24. Adults (aged 18+) using any of the six services were included. A multinomial logit regression model was used to estimate the average marginal effects for age, sex, ethnicity, and area-level deprivation, with robust patient-level standard errors and year fixed-effects.

**Results:** Within this population using any crisis-related services, adjusted analysis showed people from several ethnic minority backgrounds were more likely to be compulsorily detained or brought to a PoS by police, and less likely to use CRHTs than White individuals, with largest disparities in compulsory admissions for Black (+15.0 percentage points [pp]) and Asian (+7.3 pp) groups. Compulsory admissions were more common among older adults and men (+1.5 pp). Crisis phonelines were less commonly used by older adults, men (-4.8 pp), and people from Black (-15.6 pp) and Asian (-4.7 pp) backgrounds. Crisis cafés showed more even demographic distributions. Although adjusted differences by deprivation were small (all <1.0 pp), over half of all users lived in the two most deprived quintiles.

**Conclusions:** Among crisis-related service users, demographic groups at greater risk of coercive care are less likely to access community crisis services. Addressing these disparities could improve access and reduce inequalities.

## 1. Introduction

Inpatient mental health admissions provide intensive support for people in crisis but are commonly reported to be distressing, restrictive, (re)traumatising, disruptive to daily life, and costly (1–4). Expanding community-based crisis services has therefore been a policy priority in England. Since the early 21^st^ century, crisis resolution and home treatment teams (CRHTs) have been the main alternative to admissions, offering rapid assessment and intensive home-based care (5). Although CRHTs are established nationwide and demonstrate some effectiveness in reducing admissions (5), demand for inpatient services remains high, compulsory admissions are rising, and there is substantial service user dissatisfaction with crisis care. Recent policy in England (6) has supported the implementation of additional community-based crisis services alongside CRHTs, including low-threshold drop-in services such as crisis cafés and phonelines – to create comprehensive, integrated crisis-care pathways (7,8). Ensuring these services reach groups disproportionately subject to compulsory detention – particularly Black people and men (9–11) – is critical. However, there is little research about patterns of use of community crisis services (7).

### Study aims

This study addresses this evidence gap by exploring demographic (age, sex, ethnicity) and socioeconomic (area-level deprivation) differences in use of inpatient and community crisis services in England. We focus on service models with available national routine data: 1) crisis phonelines; 2) crisis cafés; 3) CRHTs; 4) voluntary admissions; 5) compulsory admissions under civil sections 2 and/or 3 of the Mental Health Act 1983 (MHA); and 6) conveyance to a place of safety (PoS) by the police under section 136 (s136) of the MHA.

## 2. Method

### 2.1. Study design and data source

This project was conducted by the National Institute for Health and Care Research Policy Research Unit in Mental Health (12). A working group comprising academic and lived experience researchers, and clinicians, met regularly throughout all stages of the project, contributing to key methodological decisions, and reviewing and interpreting findings.

We conducted a retrospective observational study using the Mental Health Services Data Set (MHSDS) version 5 between financial years 2021/22 and 2023/24. The MHSDS is a pseudonymised administrative database capturing information on anyone in contact with secondary mental health services funded by the NHS in England (13). As an activity-based dataset, it offers little information on care quality or experience. It comprises multiple linked sub-datasets covering patient demographics, referrals, care contacts, inpatient care, and legal status under the MHA, enabling longitudinal tracking of service use across settings.

The MHSDS provides rich service-level information but is an evolving dataset in which completeness and quality of data vary across fields. Some fields are non-mandatory, and some services are inconsistently coded, limiting the ability to generate reliable national estimates for all types of care. One of our aims was to assess what can feasibly be reported from the MHSDS regarding crisis care. NHS England monitors MHSDS data quality, which has improved over time (13).

### 2.2. Study population, outcome, and exposures

Our study population included all adults aged 18+ who had contact with at least one of the following six crisis-related service types during the study period:

1. **Crisis phonelines:** Telephone services providing access to crisis care across a catchment area.
2. **Crisis cafés:** Community drop-in services offering informal, non-clinical crisis support as an alternative to emergency departments.
3. **CRHTs:** Teams providing rapid assessment and intensive home-based treatment.
4. **Inpatient services – voluntary admissions:** Hospital admissions consented to by the patient.
5. **Inpatient services – compulsory admissions**: Hospital admissions for individuals detained without their consent under civil sections 2 (up to 28 days for assessment) and/or 3 (up to 6 months, renewable, for treatment) of the MHA, which governs compulsory mental health care in England.
6. **s136 PoS detentions:** Being brought from a public place to a PoS (usually a hospital or care facility) by police for assessment under s136 of the MHA.

All these services were identified using MHSDS service type codes. Other crisis-related services available in some areas in England (walk-in crisis assessment units, psychiatric decision units, acute day services, and crisis houses) were excluded due to insufficient available data. See Appendix A for detailed descriptions of each included and excluded service type.

We considered service use regardless of the provider and did not consider the intensity of utilisation.

We excluded individuals with dementia using the mental health care organic supercluster (14), as their crisis-care pathways and service needs differ from adults using general mental health crisis services (15). Care clusters classify patients into groups based on clinical presentation and care needs.

Compulsory admissions under section 2 and/or 3 were identified using MHA section codes and inpatient records. To reduce misclassification, we restricted these to admissions starting within 30 days of the relevant MHA legal status start date, since some of these compulsory admissions are transfers which might justify the time gap between the MHA legal start date and the admission date. We also classified admissions that began as voluntary, but where patients were subsequently detained during the same episode, as compulsory. Voluntary admissions were all admissions not classified as detentions under Part II (sections 2 and 3) and Part III (sections 35, 36, 37, 38, 44, 45A, 46, 47, and 48) of the MHA. This approach was agreed with NHS England analysts.

Individuals brought to a PoS by the police under s136 of the MHA were identified through the MHA legal status records. We only considered detentions occurring during the study period.

Our outcome was a categorical variable indicating type of crisis care used. Individuals using multiple service types were assigned separate records for each.

MHSDS has information on patients’ age (which we measured in 5-year bands), sex, and ethnicity. Ethnicity was categorised into five groups based on the 2021 Census, to avoid small subgroups, which would limit regression robustness. Socioeconomic status was proxied by the index of multiple deprivation (IMD) quintile measured at the patient’s lower layer super output area (LSOA) of residence (16). Variables such as diagnosis, care cluster, accommodation status, employment, and marital status were excluded due to extensive missingness.

### 2.3. Statistical analyses

To assess differences in patients’ demographic and socio-economic factors across the six crisis-related service types, we estimated a multinomial logit regression model with patient-level robust standard errors and year fixed-effects to account for time-varying unobserved heterogeneity. Covariates were age group, sex, ethnicity, and IMD quintile. For ease of interpretation, we computed and reported the average marginal effects (AMEs), including 95% confidence intervals (95%CI), reflecting the probability of attending each service group, for each demographic and socio-economic characteristic, while adjusting for the others in the model.

To explore how estimates changed when considering factors individually, we ran separate univariate models for age, sex, ethnicity, and IMD quintile, including year fixed-effects and patient-level robust standard errors. This allowed us to examine the extent to which associations were explained by other demographic or socio-economic variables.

All statistical analyses were conducted using Stata 18 (StataCorp LLC, College Station, TX).

### 2.4. Ethics statement

The project uses pseudonymised secondary data, and results are reported at aggregated levels, with small numbers suppressed in line with NHS England guidance. The project does not involve direct contact with patients, and patients are not identifiable from the dataset. The data were subject to the NHS National Data Opt-Out, which was applied by the data provider (17). Analyses were conducted in secure environments in line with data sharing and confidentiality protocols. Individual service user consent was not required.

## 3. Results

### 3.1. Descriptive Statistics

Table 1 summarises the socio-economic and demographic characteristics of patients accessing one of the six crisis-related services between 2021/22 and 2023/24. During the study period, around 1.4 million adults had contact with at least one of these services (80% attended only one service). The most frequently accessed services were CRHTs (used by 42% of patients), followed by crisis phonelines (38%). In contrast, only 3% of patients were brought to a PoS by the police under s136 of the MHA, and just 1% accessed crisis cafés.

**Table 1.** Descriptive statistics of the acute/crisis service users between 2021/22 – 2023/24.

|  | Crisis phonelines<br>N (%) | Crisis cafés<br>N (%) | CRHTs<br>N (%) | Voluntary admissions<br>N (%) | Compulsory<br>admissions (sections 2<br>and 3, MHA)<br>N (%) | Brought to a place of<br>safety (section 136,<br>MHA)<br>N (%) |
| --- | --- | --- | --- | --- | --- | --- |
|  | N=537,870 | N=15,000 | N=592,948 | N=96,075 | N=128,825 | N=38,386 |
| **Age (mean years)** | 42.05 | 39.17 | 42.89 | 42.94 | 45.37 | 36.13 |
| 18-22 | 69,211 (12.9%) | 1,972 (13.1%) | 67,572 (11.4%) | 9,097 (9.5%) | 11,034 (8.6%) | 5,329 (13.9%) |
| 23-27 | 66,496 (12.4%) | 2,139 (14.3%) | 68,947 (11.6%) | 10,881 (11.3%) | 13,972 (10.8%) | 5,950 (15.5%) |
| 28-32 | 64,947 (12.1%) | 1,835 (12.2%) | 69,384 (11.7%) | 11,822 (12.3%) | 13,910 (10.8%) | 6,046 (15.8%) |
| 33-37 | 59,704 (11.1%) | 1,609 (10.7%) | 66,071 (11.1%) | 11,628 (12.1%) | 13,634 (10.6%) | 5,631 (14.7%) |
| 38-42 | 50,543 (9.4%) | 1,429 (9.5%) | 57,404 (9.7%) | 10,078 (10.5%) | 12,051 (9.4%) | 4,464 (11.6%) |
| 43-47 | 41,810 (7.8%) | 1,265 (8.4%) | 47,983 (8.1%) | 8,032 (8.4%) | 10,365 (8.0%) | 3,444 (9.0%) |
| 48-52 | 41,393 (7.7%) | 1,278 (8.5%) | 48,103 (8.1%) | 7,672 (8.0%) | 10,130 (7.9%) | 2,803 (7.3%) |
| 53-57 | 37,732 (7.0%) | 1,203 (8.0%) | 43,746 (7.4%) | 6,867 (7.1%) | 9,615 (7.5%) | 2,191 (5.7%) |
| 58-62 | 28,980 (5.4%) | 899 (6.0%) | 34,322 (5.8%) | 5,393 (5.6%) | 8,630 (6.7%) | 1,343 (3.5%) |
| 63-67 | 17,718 (3.3%) | 511 (3.4%) | 23,377 (3.9%) | 4,079 (4.2%) | 6,646 (5.2%) | 622 (1.6%) |
| Above 67 | 56,653 (10.5%) | 532 (3.5%) | 65,245 (11.0%) | 10,220 (10.6%) | 18,745 (14.6%) | 508 (1.3%) |
| Missing | 2,683 (0.5%) | 328 (2.2%) | 794 (0.1%) | 306 (0.3%) | 93 (0.1%) | 55 (0.1%) |
| **Sex** | 69,211 (12.9%) | 1,972 (13.1%) | 67,572 (11.4%) | 9,097 (9.5%) | 11,034 (8.6%) | 5,329 (13.9%) |
| Female | 295,006 (54.8%) | 7,885 (52.6%) | 303,280 (51.1%) | 45,414 (47.3%) | 61,437 (47.7%) | 15,736 (41.0%) |
| Male | 235,428 (43.8%) | 6,437 (42.9%) | 282,042 (47.6%) | 49,463 (51.5%) | 66,153 (51.4%) | 22,503 (58.6%) |
| Missing | 7,436 (1.4%) | 678 (4.5%) | 7,626 (1.3%) | 1,198 (1.2%) | 1,235 (1.0%) | 147 (0.4%) |
| **Ethnicity** |  |  |  |  |  |  |
| Asian or Asian British | 23,965 (4.5%) | 370 (2.5%) | 31,371 (5.3%) | 5,309 (5.5%) | 11,546 (9.0%) | 2,432 (6.3%) |
| Black, Black British, Caribbean or African | 13,563 (2.5%) | 353 (2.4%) | 27,342 (4.6%) | 5,658 (5.9%) | 14,793 (11.5%) | 3,130 (8.2%) |
| Mixed/Multiple ethnic groups | 17,302 (3.2%) | 1,210 (8.1%) | 16,992 (2.9%) | 3,365 (3.5%) | 4,856 (3.8%) | 1,506 (3.9%) |
| Other ethnic group | 15,510 (2.9%) | 456 (3.0%) | 16,604 (2.8%) | 2,441 (2.5%) | 4,954 (3.8%) | 1,291 (3.4%) |
| White | 356,466 (66.3%) | 9,557 (63.7%) | 425,951 (71.8%) | 73,141 (76.1%) | 84,842 (65.9%) | 26,778 (69.8%) |
| Missing | 111,064 (20.6%) | 3,054 (20.4%) | 74,688 (12.6%) | 6,161 (6.4%) | 7,834 (6.1%) | 3,249 (8.5%) |
| **Index of Multiple Deprivation (quintiles)** |  |  |  |  |  |  |
| Least | 60,008 (11.2%) | 1,855 (12.4%) | 63,399 (10.7%) | 9,736 (10.1%) | 13,021 (10.1%) | 3,082 (8.0%) |
| 2 | 79,315 (14.7%) | 2,114 (14.1%) | 82,090 (13.8%) | 13,043 (13.6%) | 16,585 (12.9%) | 4,304 (11.2%) |
| 3 | 96,409 (17.9%) | 2,216 (14.8%) | 102,981 (17.4%) | 16,380 (17.0%) | 22,372 (17.4%) | 6,409 (16.7%) |
| 4 | 124,016 (23.1%) | 3,581 (23.9%) | 137,263 (23.1%) | 22,833 (23.8%) | 32,170 (25.0%) | 9,328 (24.3%) |
| Most | 170,798 (31.8%) | 4,441 (29.6%) | 195,839 (33.0%) | 31,285 (32.6%) | 40,931 (31.8%) | 14,047 (36.6%) |
| Missing | 7,324 (1.4%) | 793 (5.3%) | 11,376 (1.9%) | 2,798 (2.9%) | 3,746 (2.9%) | 1,216 (3.2%) |
| Number of patients in 2021/22 | 186,735 | 4,831 | 203,542 | 35,408 | 41,781 | 14,362 |
| Number of patients in 2022/23 | 173,920 | 4,825 | 198,391 | 30,991 | 42,481 | 12,310 |
| Number of patients in 2023/24 | 177,215 | 5,344 | 191,015 | 29,676 | 44,563 | 11,714 |

CRHTs: Crisis Resolution and Home Treatment Teams; MHA: Mental Health Act.

Over the three-year period, we observed a reduction in the number of patients accessing several crisis services: crisis phonelines (-5%), CRHTs (-6%), voluntary admissions (-16%), and being brought to a PoS by the police (-18%). Conversely, there was an increase in the number of patients accessing crisis cafés (+11%) and being compulsorily admitted under sections 2 and/or 3 of the MHA (+7%).

#### Age

The average age of patients varied by service type. Patients accessing crisis phonelines and CRHTs, as well as those voluntarily admitted, had a mean age of around 42-43 years. Patients attending crisis cafés were younger (mean age 39), whereas those admitted compulsorily were older (mean age 45). The youngest group (mean age 36), comprised individuals brought to a PoS by the police under s136 of the MHA.

Age group distributions showed that individuals aged 18-32 (the three youngest age bands) were the most represented across all service groups, especially among those brought to a PoS by the police (45%) and those attending crisis cafés (40%). For compulsory admissions, the proportion declined only gradually with age, with those aged 53-57 just 1 percentage point lower than those aged 18-22.

#### Sex

Sex distribution varied across services. Most patients accessing crisis phonelines (55%), crisis cafés (53%), and CRHTs (51%) were female. In contrast, patients voluntarily (52%) or compulsorily admitted (51%), and those brought to a PoS by the police (59%) were more likely to be male.

#### Ethnicity

White patients represented the majority in all six crisis-related service types, accounting for over 60% in each. Percentages should be interpreted with caution, however, as the proportion of missing ethnicity data was higher for crisis phonelines (21%), crisis cafés (21%), and CRHTs (13%), compared with around 6-8% in the remaining services. Among patients accessing crisis phonelines and CRHTs, the second most common ethnic group was Asian or Asian British (around 5%). For crisis cafés, the second largest group was individuals from Mixed/Multiple ethnic groups (8%). Among those admitted voluntarily, compulsorily, and those brought to a PoS by the police, the second most prevalent group was Black, Black British, Caribbean or African, comprising 6%, 12%, and 8% of patients, respectively.

For context, in the 2021 Census for England, 81.0% of the population identified as White, 9.6% as Asian or Asian British, 4.2% as Black, Black British, Caribbean or African, 3.0% as Mixed/Multiple ethnic groups, and 2.2% as Other ethnic groups (18).

#### Index of Multiple Deprivation (IMD)

Socioeconomic patterns showed a gradient, with increasing proportions of patients living in more deprived areas across all service groups. 30% or more of patients in each service group lived in areas ranked in the most deprived quintile compared to 8%-12% in the least deprived quintile.

### 3.2. Regression results

Table 2 presents the average marginal effects (AMEs) and 95% confidence intervals (95%CI) derived from the multinomial logit regression model. These AMEs indicate the change in the probability of a patient accessing a particular crisis-related service type, relative to the reference category, associated with each socio-demographic characteristic (i.e. age group, sex, ethnicity, and IMD), while controlling for the other covariates in the model. In Table 2, the AMEs are reported in their original scale (proportions). Figures 1 and 2 visually depict these effects multiplied by 100, expressed in percentage points (pp) for ease of interpretation. Figure 1 displays results for age groups (left-hand side) and sex (right-hand side). Figure 2 presents estimates for ethnicity (top graph) and IMD quintiles (bottom graph). To aid interpretation, we also estimated univariate models including one characteristic at a time. These results are presented in Appendix B.

**Figure 1.**
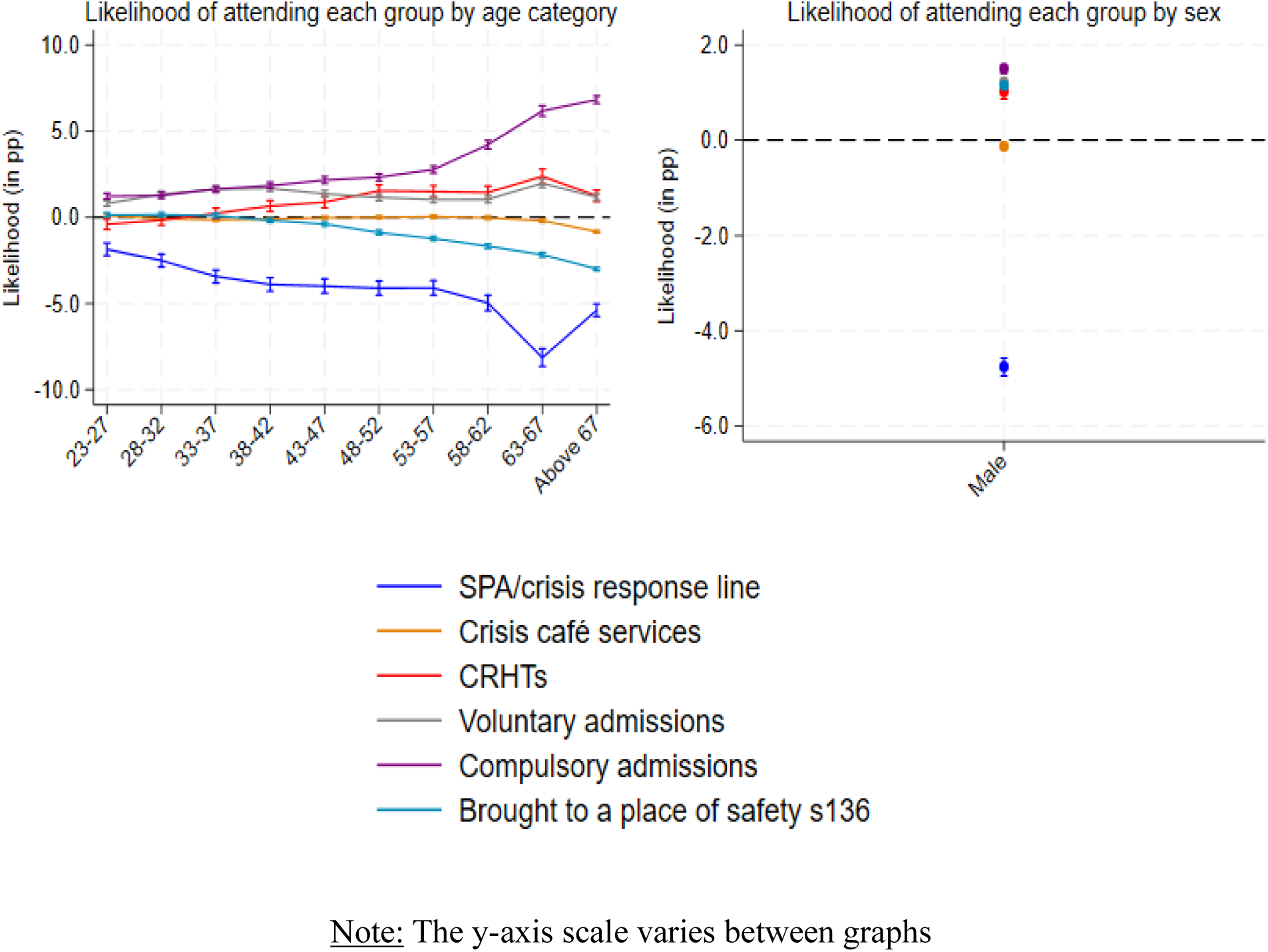
Average Marginal Effects (in percentage points) for age groups and sex among acute/crisis service users. <u>Note:</u> The y-axis scale varies between graphs

**Figure 2.**
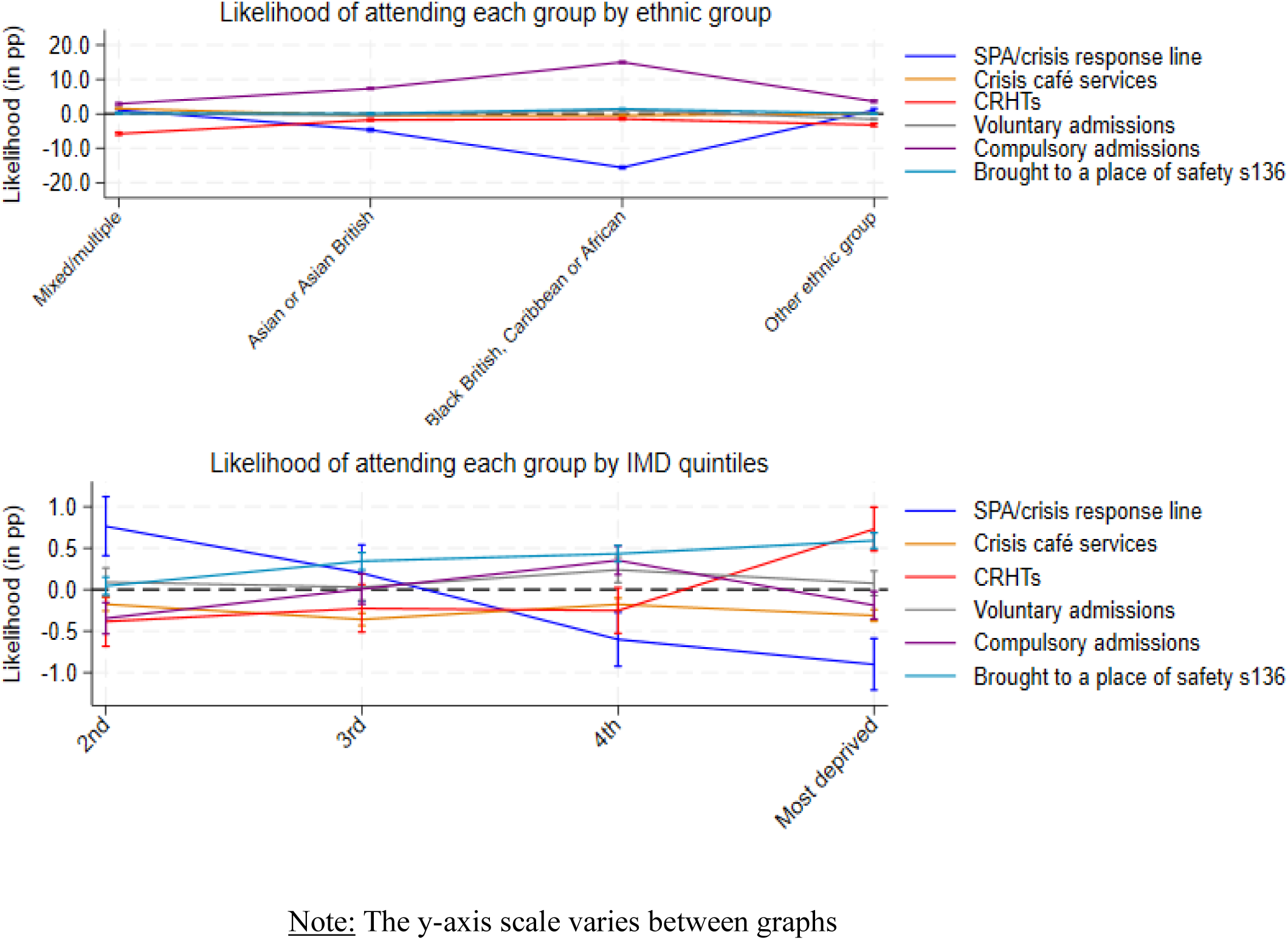
Average Marginal Effects (in percentage points) for ethnicity and IMD quintiles among acute/crisis service users. <u>Note:</u> The y-axis scale varies between graphs

**Table 2.**
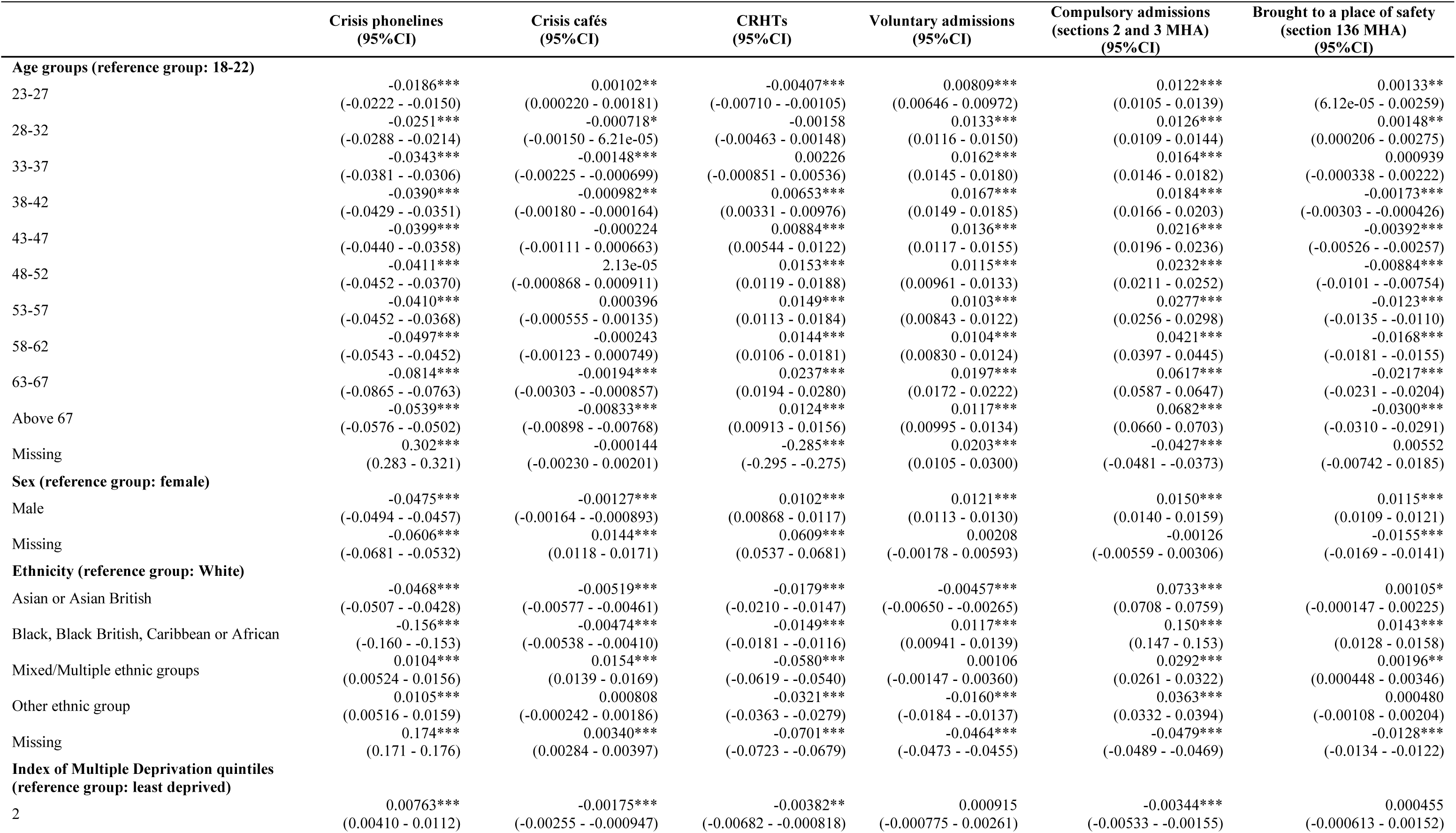
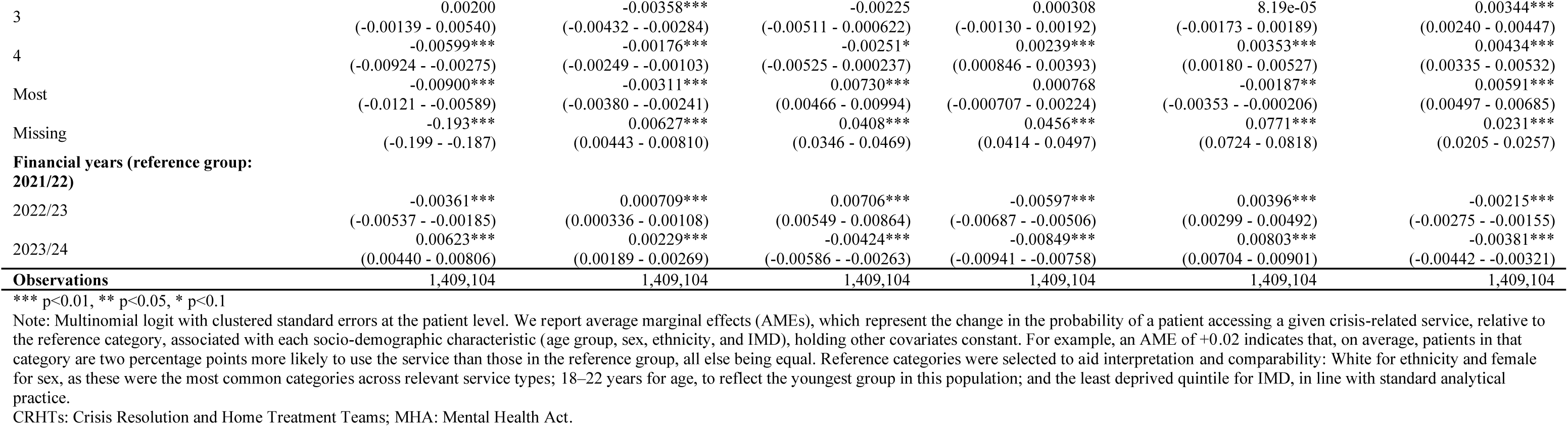
Regression results: average marginal effects among acute/crisis service users.

#### Age group

Age was significantly associated with variation in service use. In all models, patients aged 18-22 were the reference group. Crisis phonelines showed the most distinct age-related patterns. All age groups were less likely to access this service compared to patients aged 18-22, with likelihood decreasing progressively with age. The most pronounced difference was observed among patients aged 63-67 who were 8.1 pp less likely to use this than those aged 18-22.

For voluntary admissions, likelihood increased progressively among patients 23-42 compared with the youngest group. Compulsory admissions were more pronounced from age 43 onwards, and patients aged 38 and over were also more likely to use CRHTs than those aged 18-22. For example, patients over 67 were 6.8 pp more likely to be compulsorily admitted and 3.0 pp less likely to be brought to a PoS by the police compared to the youngest group. The likelihood of attending crisis cafés did not differ markedly across most age groups, although patients aged 63 and above were significantly less likely to attend than 18–22-year-olds.

#### Sex

Males were less likely to use crisis phonelines and crisis cafés than females by 4.8 pp and 0.1 pp, respectively. While the effect for crisis cafés was statistically significant, it was substantively small, suggesting minimal differences between male and female patients. For the remaining services, male patients were more likely to access them than females. The AMEs for CRHTs, voluntary admissions, and being brought to a PoS by the police were similar (with overlapping 95%CI) indicating no substantial differences between services. However, for compulsory admissions, male patients were 1.5 pp more likely to be admitted (see Figure 1).

#### Ethnicity

Ethnic disparities were evident in several service types. Compared with White patients, individuals from all other ethnic backgrounds were more likely to be compulsorily admitted. This difference was most pronounced among Black patients, who were 15.0 pp more likely to be compulsorily admitted, and among Asian ethnic groups, who were 7.3 pp more likely.

All ethnic minority groups were less likely to access CRHTs than White patients, particularly patients of Mixed/Multiple ethnic backgrounds, who were 5.8 pp less likely.

Asian and Black patients were significantly less likely to use crisis phonelines than White patients by 4.7 pp and 15.6 pp, respectively. Conversely, patients from Mixed/Multiple, and Other ethnic backgrounds were more likely to use crisis phonelines, although these differences were small (1.0 pp and 1.1 pp, respectively).

Differences in the use of crisis cafés by ethnicity were generally small but statistically significant, except for the Other ethnic group, which was not statistically significant.

Asian and Other ethnic groups were less likely to be voluntarily admitted (0.5 pp and 1.6 pp, respectively), while Black patients were 1.2 pp more likely to be voluntarily admitted than White patients. There was no statistically significant difference in the likelihood of voluntary admission for patients from Mixed/Multiple ethnic backgrounds.

Black patients were also 1.4 pp more likely to be brought to a PoS by the police than White patients. Patients from Mixed/Multiple ethnic backgrounds were 0.2 pp more likely to experience this outcome than White patients, but differences for the remaining ethnic groups were not statistically significant.

#### Index of Multiple Deprivation (IMD)

Differences in crisis service use by area-level deprivation (IMD quintiles) were small (all <1.0 pp). Compared with the least deprived quintile, individuals in the most deprived quintiles were slightly more likely to use CRHTs and be brought to a PoS by police, and slightly less likely to use crisis phonelines or crisis cafes. Crisis phoneline use consistently declined, and being brought to a PoS by police consistently increased, with increasing deprivation. Patterns for other service types across quintiles were less consistent (full results in Table 2).

#### Univariate analyses

Results from univariate models showed patterns consistent with the fully-adjusted analysis. Strong univariate associations, including the age gradient in crisis phoneline use, the higher likelihood of compulsory admission among Black patients, and sex differences in crisis phoneline use and compulsory admissions remained evident after adjusting for other characteristics. Smaller univariate associations, particularly for voluntary admissions across some ethnic and deprivation groups, were attenuated once covariates were included in the fully adjusted analysis (see Appendix B for full results).

## 4. Discussion

This study explored socio-demographic differences in the use of six NHS-funded mental health crisis-related services in England, using a national administrative dataset. Over half of users lived in the two most deprived quintiles, highlighting the concentration of crisis-service use in more deprived areas. Among those accessing crisis-related services, adjusted analyses showed clear disparities in service use for ethnicity, sex, and age, while adjusted differences by area-level deprivation were small, indicating limited variation in which crisis services people use once in contact with crisis care.

Ethnic inequalities were most pronounced: all minority ethnic groups were more likely than White individuals to be compulsorily admitted rather than using other crisis-related services, with the largest disparities for Black, then Asian groups. Minority ethnic groups were also less likely to access CRHTs. Use of lower-intensity services such as crisis cafés and crisis phonelines was substantially lower among Black groups and moderately lower among Asian groups, compared to White individuals. These findings extend previous evidence of disproportionately high compulsory detention rates among Black and Asian ethnic groups, and that Black groups are more likely to encounter mental health services via the criminal justice system (9,10,19–21). Our findings show these ethnic inequalities persist across a broad range of crisis services, with relatively low use of community crisis services in comparison to compulsory detention rates. Contributing factors could include: racist stereotypes influencing services offered and decisions about detention, cultural preferences and understanding, stigma associated with use of mental health services, differences in help-seeking, higher prevalence of psychosis, lack of culturally appropriate care, or mistrust in services (10,11,22–24).

Sex and age differences were also evident but smaller. Men accessed crisis phonelines and crisis cafés proportionately less than women but were more likely to use CRHTs, be voluntarily or compulsorily admitted, or brought to a PoS by police. This aligns with existing evidence of higher detention rates (9,25), and lower mental health treatment-seeking rates (26–28) among men. Possible explanations include sex differences in distress expression (e.g. displaying more externalising behaviours such as aggression or substance use), pressure to conform to traditional masculine stereotypes (e.g., stoicism, self-reliance, invulnerability), differences in coping strategies, help-seeking, emotional literacy, stigma, clinician bias (e.g., overlooking distress), and service acceptability (29–32).

Older crisis-related service users were less likely to use crisis phonelines or be taken to a PoS by police, but more likely to use CRHTs, or be voluntarily or compulsorily admitted. Barriers to accessing lower-intensity community crisis services for older adults could relate to unawareness of services, digital exclusion, vision, hearing or physical disability, stigma, mistrust of services, and acceptability, and failure to recognise a need for support (33,34). Notably, compulsory detention rates remained relatively high across older age groups, highlighting that initiatives to reduce detentions should not just target young adults – crisis-response policies and interventions that specifically consider the needs of older adults are needed (33).

Most crisis service users lived in more deprived areas, with the proportion using each service type increasing with deprivation. However, differences in the types of services accessed by area-level deprivation were very small (all <1.0 pp) and notably smaller than differences observed for other demographic characteristics, particularly ethnicity. This suggests area-level deprivation influences whether people use crisis services but has little impact on which crisis services they use. These findings are broadly consistent with evidence linking higher levels of area deprivation to higher rates of mental health difficulties (35–37), urgent and emergency care use (38) and compulsory detentions (9).

Altogether, the findings generally suggest that groups with higher compulsory admission rates – including Black and Asian patients, men, and older adults – also tend to engage less with lower-intensity services such as crisis phonelines when experiencing a crisis. Differences for Black patients were substantially larger than those for any other ethnic group or demographic characteristic. As this is an observational study, we cannot establish causality, but the findings suggest associations between lower engagement with lower-intensity community crisis services and higher rates of coercive care, which warrants further investigation. Although some estimated differences were small in percentage points terms, applied to the large population using crisis services nationally, they represent substantial absolute numbers and remain of policy relevance.

### Strengths and limitations

A key strength of this study is the use of a large, national administrative dataset (MHSDS), which allowed comparisons across multiple crisis-related services in England. Additionally, lived experience researchers within the study team shaped the research questions and interpretation of findings, ensuring the study reflected service users’ and carers’ perspectives. Multivariate analysis allowed examination of the combined influence of demographic and socioeconomic factors on service use, helping to identify disparities among people accessing crisis care. This addresses an important evidence gap around how community-based crisis models are being used in practice.

Limitations include limited available data in MHSDS on some crisis service types (e.g., crisis houses, acute day services, walk-in crisis assessment units), which we therefore could not include. This is likely due to challenges in how these services are reported or classified in the dataset and inconsistent reporting across areas where they are available. Second, limited clinical data (e.g. diagnosis or care cluster) prevented examining how service use differences relate to clinical need or severity. Third, MHSDS lacks some relevant demographic characteristics (e.g. sexual orientation, disability) related to mental health. Fourth, ethnicity was measured using broad categories, which may obscure within-group variation, but was necessary to obtain reliable regression estimates given the small numbers in some subcategories. Finally, IMD-based deprivation reflects area-level rather than individual socioeconomic circumstances. Together, these limitations constrain how fully we could capture the complexity of sociodemographic and clinical factors’ influences on crisis service use.

### Future research, practice, and policy

Mixed methods research involving service users, families/carers, and staff is needed to better understand the mechanisms driving these differences, and to establish guidance for culturally aware, inclusive crisis care practices which can reduce inequalities in use and experiences of services. This is especially important for groups who tend to use high-intensity inpatient services rather than community crisis services – including older adults, men, and people from Black ethnic groups. For example, research could clarify why rates of voluntary and compulsory detentions remain high across older age groups. Our study highlights challenges in community crisis care access for people from Black ethnic groups, who are much more likely than others to be coerced and less likely to use community alternatives. Addressing the reasons for this should be a priority for research, policy and practice; supported by initiatives like the Patient and Carer Race Equality Framework (39).

Commissioners and service providers should demographically monitor local service use, meaningfully involve under-represented groups to understand barriers and inform service development, and implement both targeted interventions and wider structural changes to improve acceptability, equity and access across the crisis-care pathway.

Crisis cafés are an emerging model (40), and our findings provide preliminary evidence that, where available, they are accessible and inclusive across demographic groups. Further research is warranted to understand their role in the mental health pathway, their effectiveness in reducing hospital admission rates and producing good outcomes, and user experience.

Observed patterns may partly reflect local service availability. National mapping of crisis care alongside improved routine data collection for under-represented services – including crisis houses, acute day services, and walk-in crisis assessment units (8), and emerging models such as mental health A&Es (6) – would allow evaluation of their reach, accessibility, and impact, supporting more equitable and evidence-based commissioning. In addition, improving the completeness of MHSDS, or linking it with other datasets, would reduce missing data and allow inclusion of other clinical and sociodemographic variables in analyses, such as diagnosis, accommodation, employment, migrant status, disability, gender identity, sexual orientation, and other protected characteristics. Analyses could also examine intersectionality by testing interaction effects and extend our work by examining intensity of crisis service use. These approaches could clarify how sociodemographic and clinical factors influence crisis service use and support efforts to reduce inequalities.

## 5. Conclusions

Using national administrative data (MHSDS), this study highlights sociodemographic patterns in NHS-funded mental health crisis service use in England. Ethnic disparities were most pronounced, particularly for Black patients, who were much more likely to be compulsorily admitted and less likely to access lower-intensity community services. Smaller but similar patterns were observed for men, and older adults. These findings highlight the need to better understand and address the factors limiting access to community crisis care and increasing the risk of compulsory admissions.

## 6. Lived experience commentary by Jude Beng and Mark Holden

We welcome this retrospective observational research study, which used the Mental Health Services Dataset (MHSDS) between 2021/22 and 2023/24.

However, our primary concerns are:

1. The higher rates of compulsory detentions for people from Black, Asian and/or Minority Ethnic (BAME) groups than for white people. Institutional, structural, systemic and in-person prejudices, stereotypes, and racism (overt and/or covert) are plausible root causes of these disparities in outcomes, and the lack of cultural, sensitive, appropriately tailored treatments.
2. The coercive care, including being brought to a place of safety by police, is more common among older adults and men. Difficult treatment experiences have discouraged most of these patients from accessing other community crisis care services, that women are more likely to use, such as crisis phone-lines, crisis cafes and Crisis Resolution and Home Treatments (CRHTs), unless they are in desperate need.

Over 1.4 million people accessed crisis services during the study period. The researchers didn’t look at whether people in mental health crises accessed services once, multiple times, or not at all, and consequently had unmet needs. Some crisis service types and demographics weren’t included because of missing data. Furthermore, we are unaware of crisis cafes available in our locale and the study didn’t explore ‘postcode lotteries’ between different regions in England either.

It’s important to consider societal and cultural contexts, when focussing on the priority outcome from this study to address these disparities in crisis care and outcomes, by changing mental health policy and practice, hence improve access and reduce clear inequalities.

Finally, this study does not involve patient interviews, whereby they shared their experiences and could give possible explanations for these deeply personal issues - there is scope for this in future research. These patients are unidentifiable from the dataset, so direct follow-up with each of them is impossible.

## Data availability statement

The data supporting the findings of this study are available from NHS England. Restrictions apply to the availability of the following datasets that were used under licence for this study. This work uses data provided by patients and collected by the NHS as part of their care and support. The Mental Health Services Data Set is copyright ©2021/22-2023/24, NHS England. Re-used with the permission of NHS England. All rights reserved.

## Data Availability

The data supporting the findings of this study are available from NHS England. Restrictions apply to the availability of the following datasets that were used under licence for this study. This work uses data provided by patients and collected by the NHS as part of their care and support. The Mental Health Services Data Set is copyright 2021/22-2023/24, NHS England. Re-used with the permission of NHS England. All rights reserved.

## Acknowledgements

We are grateful for the support from York Data Safe Heaven and Research Computing teams. This project was conducted by the National Institute for Health and Care Research (NIHR) Policy Research Unit in Mental Health (MHPRU), based at University College London, King’s College London, and University of York. The NIHR MHPRU conducts research in response to policymaker need (e.g., in the Department of Health and Social Care or NHS England).

## Authors contribution

BLE, RJ, SJ, and MAM formulated the research question. BLE, RJ, SJ, and MAM designed the study. MAM analysed the data. MAM and JG wrote the first draft of the manuscript. JB and MH wrote the lived experience commentary. BLE, RJ, AB, BC, RRO, MH, JB, AG, PN, DO, JEC, KT, MW, SJ, and AS critically reviewed the manuscript. All authors approved the final version to be published.

## Appendices

**Appendix A:** More detailed crisis-related service definitions (based on NHS digital definitions, available here: https://digital.nhs.uk/data-and-information/data-collections-and-data-sets/data-sets/mental-health-services-data-set/submit-data/urgent-and-emergency-mental-health-care-pathways/appendix)

### Included services

- <u>Crisis phoneline:</u> Teams where the primary function is operating a Single Point of Access service, including a mental health enquiry and/or crisis response line. They provide immediate telephone or online support, assessment, and signposting for people experiencing a mental health crisis.
- <u>Crisis cafés:</u> Safe, homely community spaces, often open in the evenings and weekends, offering practical and emotional support to people in crisis as an alternative to A&E. They focus on reducing immediate crisis through safety planning, drawing on strengths and coping skills, and may also provide referrals and signposting to other services. They may include a 24-hour crisis support line.
- <u>Crisis Resolution and Home Treatment Teams (CRHTs):</u> Multidisciplinary teams providing urgent mental health assessments, gatekeeping inpatient admissions, providing intensive home treatment as an alternative to admission, and facilitating early discharge from inpatient care.
- <u>Inpatient services - voluntary admissions:</u> Hospital admissions where patients consent to stay and receive treatment on mental health wards without being compulsorily detained under the Mental Health Act 1983.
- <u>Inpatient services - compulsory admissions under</u> sections 2 and 3 <u>of the Mental Health Act (1983):</u> Hospital detentions for assessment (section 2) or treatment (section 3) of individuals without their consent.
- <u>Police detentions under</u> section 136 <u>of the MHA (place of safety):</u> Police can detain individuals they believe to have a mental disorder, and who may cause harm to themselves or another, in a public place and take them to a designated place of safety – usually a hospital or care facility – for assessment. Police stations should only be used in exceptional circumstances.

### Excluded services

- <u>Walk-in crisis assessment units:</u> Open-access NHS facilities where people in mental health crisis can access immediate support and assessment. Individuals can self-refer or be brought by partners such as police or ambulance without prior NHS referral. They are mainly staffed by mental health nurses and other qualified professionals. These units are sometimes considered the mental health equivalent of A&E.
- <u>Psychiatric decision units:</u> Acute mental health assessment units providing short-term support (usually up to 48-72 hours) and enhanced assessment for people in crisis, often referred from A&E or urgent mental health services. These units offer respite and time for ongoing assessment and treatment planning, aiming to potentially reduce unnecessary inpatient admissions by supporting tailored, community-based care.
- <u>Acute day services:</u> Services providing assessment and treatment to people experiencing a mental health crisis as an alternative to inpatient admission or to reduce inpatient stay length. Treatment provided is comparable to inpatient care and can be provided as part of an acute hospital unit or as a separate unit. Some also support relapse prevention or recovery for individuals not requiring intensive home treatment.
- <u>Crisis houses:</u> Community-based residential services offering short-term clinical and social support for people in mental health crisis as an alternative to hospital admission. Some provide specialist support to specific populations, such as women. Typically delivered in partnership with voluntary or social care organisations, they provide a safe, supportive environment where people can recover while maintaining independence. Some are staffed mainly by voluntary sector workers and/or peer support workers, some mainly by clinical staff, and some by a mix.

**Appendix B:** Results of univariate analyses

**Figure S1.**
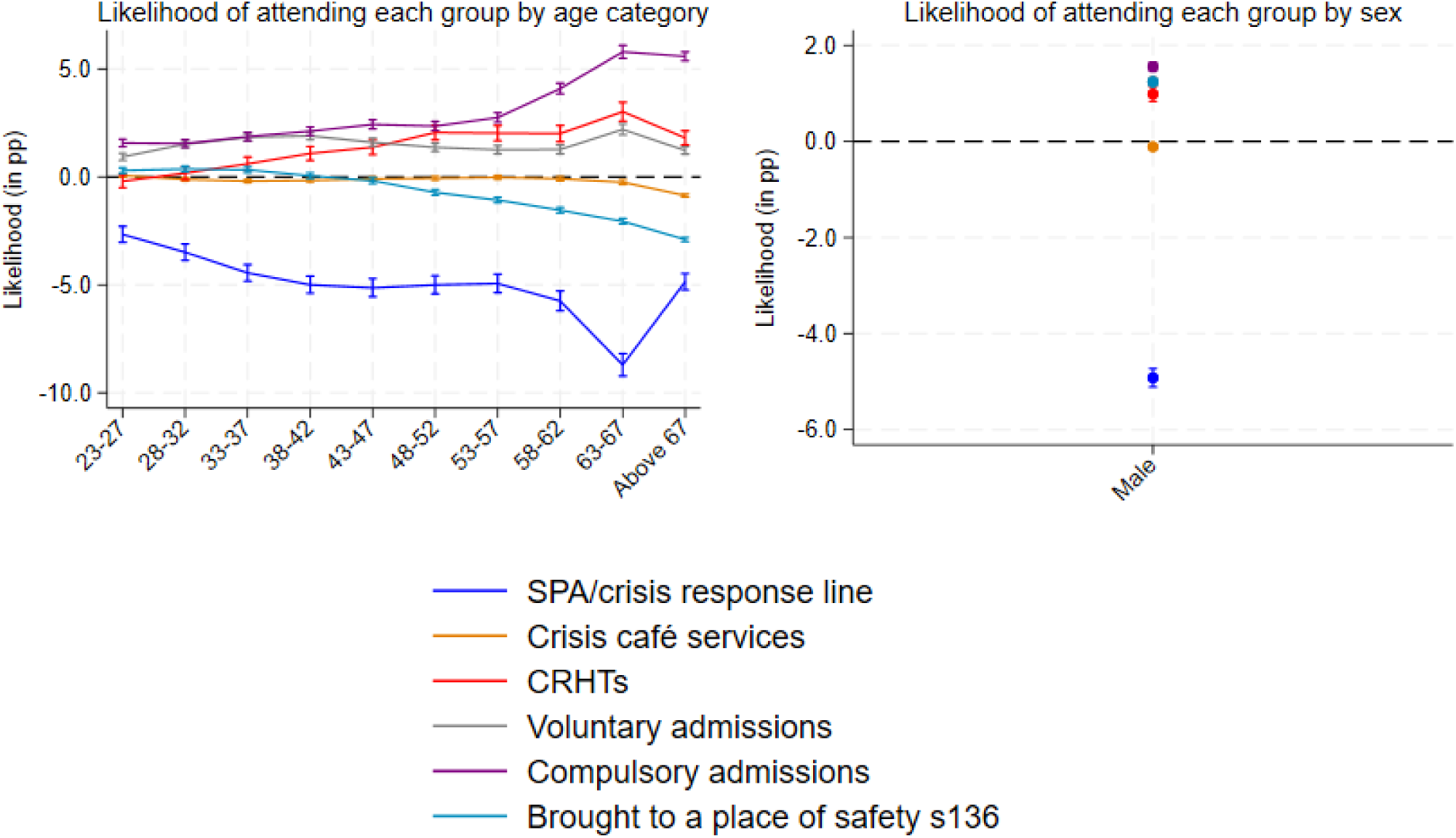
Univariate analysis: Average Marginal Effects (in percentage points) for age groups and sex among acute/crisis service users. <u>Note:</u> The y-axis scale varies between graphs

**Figure S2.**
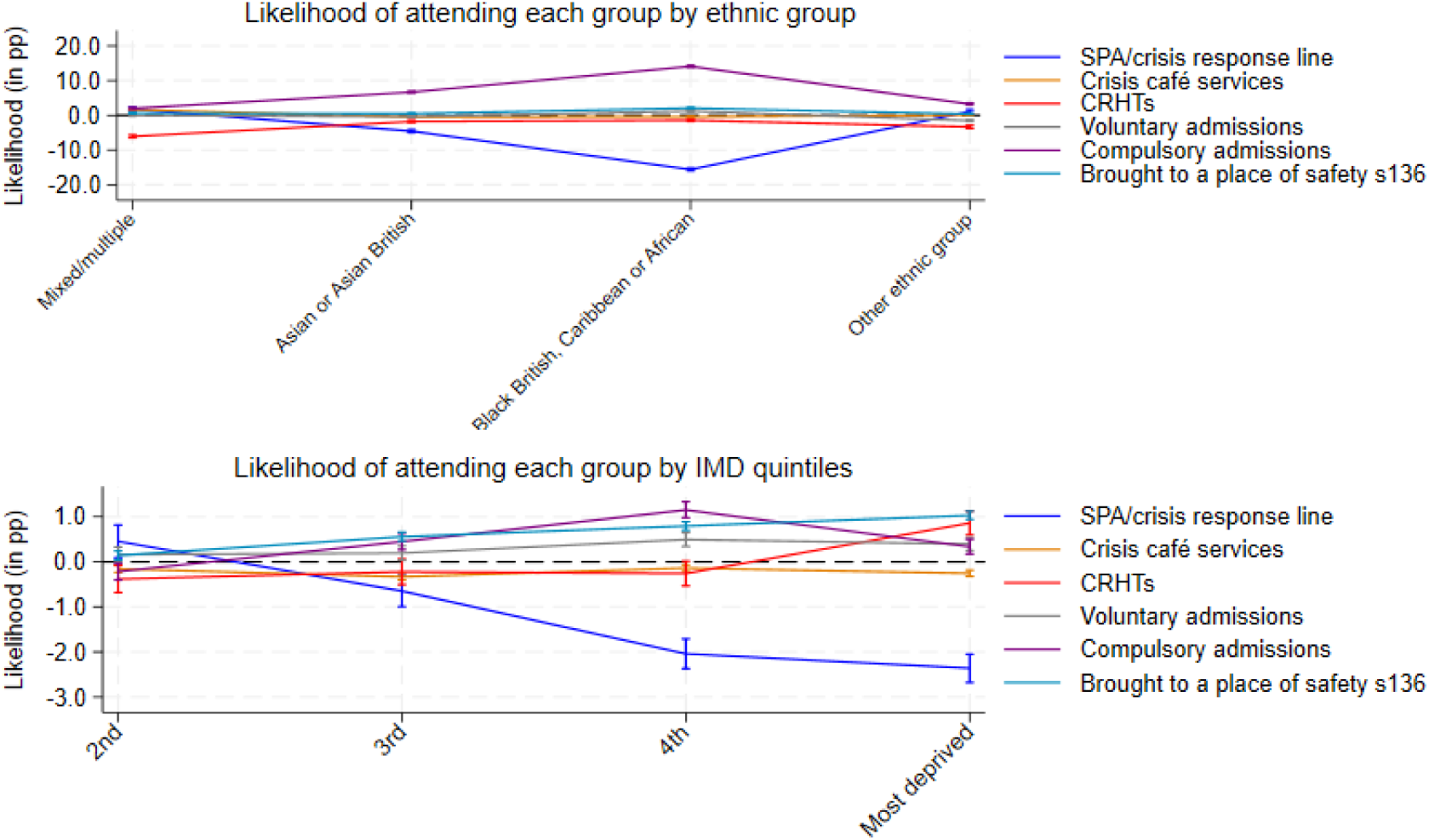
Univariate analysis: Average Marginal Effects (in percentage points) for ethnicity and IMD quintiles among acute/crisis service users. <u>Note:</u> The y-axis scale varies between graphs

**Table S1.** Regression results (univariate analysis) for age: average marginal effects among acute/crisis service users.

|  | Crisis phonelines | Crisis cafés | CRHTs | Voluntary admissions | Compulsory admissions (sections 2 and 3, MHA) | Brought to a place of safety (section 136, MHA) |
| --- | --- | --- | --- | --- | --- | --- |
| **Age groups (reference group: 18-22)** |  |  |  |  |  |  |
| 23-27 | -0.0266***<br>(-0.0304 - -0.0229) | 0.000659<br>(-0.000145 - 0.00146) | -0.00198<br>(-0.00500 - 0.00104) | 0.00930***<br>(0.00769 - 0.0109) | 0.0157***<br>(0.0139 - 0.0175) | 0.00294***<br>(0.00169 - 0.00419) |
| 28-32 | -0.0349***<br>(-0.0387 - -0.0311) | -0.00115***<br>(-0.00193 - -0.000362) | 0.00175<br>(-0.00130 - 0.00479) | 0.0151***<br>(0.0135 - 0.0168) | 0.0155***<br>(0.0137 - 0.0173) | 0.00365***<br>(0.00238 - 0.00492) |
| 33-37 | -0.0445***<br>(-0.0483 - -0.0406) | -0.00194***<br>(-0.00272 - -0.00116) | 0.00609***<br>(0.00299 - 0.00919) | 0.0183***<br>(0.0166 - 0.0201) | 0.0187***<br>(0.0168 - 0.0205) | 0.00329***<br>(0.00201 - 0.00457) |
| 38-42 | -0.0499***<br>(-0.0539 - -0.0460) | -0.00159***<br>(-0.00241 - -0.000780) | 0.0108***<br>(0.00763 - 0.0141) | 0.0190***<br>(0.0172 - 0.0208) | 0.0212***<br>(0.0192 - 0.0231) | 0.000536<br>(-0.000766 - 0.00184) |
| 43-47 | -0.0514***<br>(-0.0555 - -0.0472) | -0.000905**<br>(-0.00178 - -2.73e-05) | 0.0137***<br>(0.0103 - 0.0171) | 0.0160***<br>(0.0141 - 0.0179) | 0.0243***<br>(0.0222 - 0.0264) | -0.00179***<br>(-0.00313 - -0.000440) |
| 48-52 | -0.0500***<br>(-0.0542 - -0.0458) | -0.000610<br>(-0.00149 - 0.000272) | 0.0205***<br>(0.0171 - 0.0240) | 0.0137***<br>(0.0118 - 0.0155) | 0.0236***<br>(0.0215 - 0.0257) | -0.00717***<br>(-0.00845 - -0.00588) |
| 53-57 | -0.0494***<br>(-0.0537 - -0.0451) | -0.000221<br>(-0.00116 - 0.000721) | 0.0203***<br>(0.0168 - 0.0238) | 0.0126***<br>(0.0106 - 0.0145) | 0.0275***<br>(0.0253 - 0.0296) | -0.0107***<br>(-0.0120 - -0.00944) |
| 58-62 | -0.0575***<br>(-0.0621 - -0.0528) | -0.000825*<br>(-0.00181 - 0.000156) | 0.0201***<br>(0.0163 - 0.0239) | 0.0127***<br>(0.0106 - 0.0148) | 0.0409***<br>(0.0384 - 0.0434) | -0.0154***<br>(-0.0167 - -0.0141) |
| 63-67 | -0.0871***<br>(-0.0922 - -0.0819) | -0.00247***<br>(-0.00354 - -0.00139) | 0.0302***<br>(0.0258 - 0.0345) | 0.0220***<br>(0.0195 - 0.0245) | 0.0579***<br>(0.0549 - 0.0609) | -0.0206***<br>(-0.0219 - -0.0193) |
| Above 67 | -0.0486***<br>(-0.0523 - -0.0448) | -0.00859***<br>(-0.00925 - -0.00793) | 0.0181***<br>(0.0148 - 0.0213) | 0.0123***<br>(0.0105 - 0.0140) | 0.0558***<br>(0.0538 - 0.0579) | -0.0290***<br>(-0.0299 - -0.0280) |
| Missing | 0.205***<br>(0.190 - 0.220) | 0.0736***<br>(0.0648 - 0.0823) | -0.225***<br>(-0.237 - -0.214) | 0.0118***<br>(0.00471 - 0.0189) | -0.0446***<br>(-0.0493 - -0.0399) | -0.0203***<br>(-0.0236 - -0.0171) |
| **Observations** | 1,409,104 | 1,409,104 | 1,409,104 | 1,409,104 | 1,409,104 | 1,409,104 |
| **Year fixed-effects** | Yes | Yes | Yes | Yes | Yes | Yes |

\*\*\* p<0.01, \*\* p<0.05, \* p<0.1
Note: Multinomial logit with clustered standard errors at the patient level. We report average marginal effects (AMEs), which represent the change in the probability of a patient accessing a given crisis-related service, relative to the reference category, associated with each socio-demographic characteristic (age group, sex, ethnicity, and IMD), holding other covariates constant. For example, an AME of +0.02 indicates that, on average, patients in that category are two percentage points more likely to use the service than those in the reference group, all else being equal.

**Table S2.**
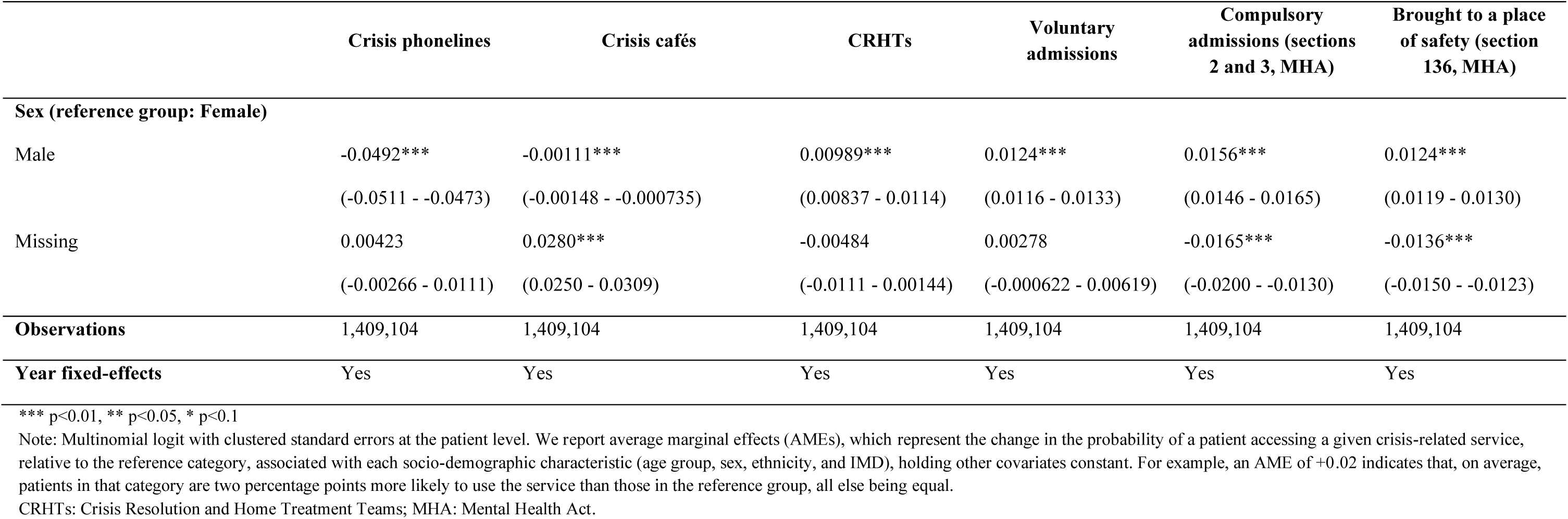
Regression results (univariate analysis) for sex: average marginal effects among acute/crisis service users.

**Table S3.** Regression results (univariate analysis) for ethnicity: average marginal effects among acute/crisis service users.

|  | Crisis phonelines | Crisis cafés | CRHTs | Voluntary admissions | Compulsory admissions (sections 2 and 3, MHA) | Brought to a place of safety (section 136, MHA) |
| --- | --- | --- | --- | --- | --- | --- |
| **Ethnicity (reference group: White)** |  |  |  |  |  |  |
| Asian or Asian British | -0.0454***<br>(-0.0494 - -0.0415) | -0.00487***<br>(-0.00545 - -0.00429) | -0.0177***<br>(-0.0208 - -0.0146) | -0.00393***<br>(-0.00586 - -0.00200) | 0.0668***<br>(0.0644 - 0.0693) | 0.00510***<br>(0.00377 - 0.00643) |
| Black, Black British, Caribbean or African | -0.156***<br>(-0.160 - -0.152) | -0.00436***<br>(-0.00501 - -0.00370) | -0.0143***<br>(-0.0176 - -0.0111) | 0.0125***<br>(0.0103 - 0.0148) | 0.141***<br>(0.138 - 0.144) | 0.0210***<br>(0.0193 - 0.0226) |
| Mixed/Multiple ethnic groups | 0.0177***<br>(0.0125 - 0.0229) | 0.0168***<br>(0.0152 - 0.0184) | -0.0607***<br>(-0.0647 - -0.0567) | 9.47e-05<br>(-0.00240 - 0.00259) | 0.0199***<br>(0.0170 - 0.0228) | 0.00621***<br>(0.00453 - 0.00789) |
| Other ethnic group | 0.0108***<br>(0.00542 - 0.0161) | 0.00116**<br>(0.000108 - 0.00222) | -0.0333***<br>(-0.0375 - -0.0291) | -0.0152***<br>(-0.0176 - -0.0129) | 0.0325***<br>(0.0294 - 0.0355) | 0.00418***<br>(0.00247 - 0.00590) |
| Missing | 0.174***<br>(0.171 - 0.177) | 0.00504***<br>(0.00446 - 0.00562) | -0.0736***<br>(-0.0757 - -0.0714) | -0.0450***<br>(-0.0459 - -0.0441) | -0.0489***<br>(-0.0499 - -0.0479) | -0.0116***<br>(-0.0123 - -0.0110) |
| **Observations** | 1,409,104 | 1,409,104 | 1,409,104 | 1,409,104 | 1,409,104 | 1,409,104 |
| **Year fixed-effects** | Yes | Yes | Yes | Yes | Yes | Yes |

\*\*\* p<0.01, \*\* p<0.05, \* p<0.1
Note: Multinomial logit with clustered standard errors at the patient level. We report average marginal effects (AMEs), which represent the change in the probability of a patient accessing a given crisis-related service, relative to the reference category, associated with each socio-demographic characteristic (age group, sex, ethnicity, and IMD), holding other covariates constant. For example, an AME of +0.02 indicates that, on average, patients in that category are two percentage points more likely to use the service than those in the reference group, all else being equal.

**Table S4.** Univariate analysis: Regression results for deprivation – average marginal effects among acute/crisis service users.

|  | Crisis phonelines | Crisis cafés | CRHTs | Voluntary admissions | Compulsory admissions (sections 2 and 3, MHA) | Brought to a place of safety (section 136, MHA) |
| --- | --- | --- | --- | --- | --- | --- |
| **IMD quintiles (reference group: least deprived)** |  |  |  |  |  |  |
| 2 | 0.00455**<br>(0.000927 - 0.00817) | -0.00157***<br>(-0.00235 - -0.000789) | -0.00382**<br>(-0.00684 - -0.000810) | 0.00161*<br>(-5.12e-05 - 0.00328) | -0.00217**<br>(-0.00402 - -0.000310) | 0.00140***<br>(0.000420 - 0.00237) |
| 3 | -0.00649***<br>(-0.00996 - -0.00302) | -0.00330***<br>(-0.00402 - -0.00258) | -0.00221<br>(-0.00509 - 0.000659) | 0.00193**<br>(0.000343 - 0.00353) | 0.00449***<br>(0.00269 - 0.00630) | 0.00557***<br>(0.00460 - 0.00654) |
| 4 | -0.0204***<br>(-0.0237 - -0.0171) | -0.00140***<br>(-0.00212 - -0.000688) | -0.00259*<br>(-0.00533 - 0.000147) | 0.00494***<br>(0.00342 - 0.00646) | 0.0115***<br>(0.00979 - 0.0133) | 0.00795***<br>(0.00702 - 0.00887) |
| Most | -0.0236***<br>(-0.0268 - -0.0205) | -0.00256***<br>(-0.00324 - -0.00188) | 0.00858***<br>(0.00594 - 0.0112) | 0.00395***<br>(0.00250 - 0.00541) | 0.00336***<br>(0.00172 - 0.00500) | 0.0103***<br>(0.00942 - 0.0112) |
| Missing | -0.129***<br>(-0.135 - -0.123) | 0.0171***<br>(0.0150 - 0.0192) | -0.00196<br>(-0.00769 - 0.00378) | 0.0374***<br>(0.0338 - 0.0411) | 0.0523***<br>(0.0482 - 0.0564) | 0.0238***<br>(0.0213 - 0.0263) |
| **Observations** | 1,409,104 | 1,409,104 | 1,409,104 | 1,409,104 | 1,409,104 | 1,409,104 |
| **Year fixed-effects** | Yes | Yes | Yes | Yes | Yes | Yes |

\*\*\* p<0.01, \*\* p<0.05, \* p<0.1
Note: Multinomial logit with clustered standard errors at the patient level. We report average marginal effects (AMEs), which represent the change in the probability of a patient accessing a given crisis-related service, relative to the reference category, associated with each socio-demographic characteristic (age group, sex, ethnicity, and IMD), holding other covariates constant. For example, an AME of +0.02 indicates that, on average, patients in that category are two percentage points more likely to use the service than those in the reference group, all else being equal.

